# LEPROSY RELAPSE AFTER POLYCHEMOTHERAPY: A SYSTEMATIC REVIEW AND META-ANALYSIS

**DOI:** 10.1101/2023.12.19.23300270

**Authors:** Fabiane Verônica da Silva, Gutembergue Santos de Sousa, Elena Alves Benevides Ferreira, Pãmela Rodrigues de Souza Silva, Juliana Akie Takahashi, Omar Ariel Espinosa, Eliane Ignotti, Roberta Pinheiro Olmo, Vilanice Alves de Araújo Püschel, Zélia Ferrreira Caçador Anastácio, Silvana Margarida Benevides Ferreira

**Author notes:** Corresponding Author (F.V.S).

## Abstract

**Objective**: To synthesize the best scientific evidence related to estimating the prevalence of leprosy relapse cases after polychemotherapy treatment. **Method:** A systematic review was conducted following the JBI methodology for systematic reviews of prevalence studies, and the reporting stage adhered to PRISMA-P, with registration No.: CRD42020177141. The inclusion criteria were adopted following the PopCoCo mnemonic (Population, Condition, Context). Population: people of both genders and any age, diagnosed with leprosy relapse, and treated with paucibacillary or multibacillary therapeutic regimes. Condition: leprosy relapse after Polychemotherapy (PCT) estimated as a proportion of cases. **Context**: studies conducted within the scope of health services. Databases used: Medline, LILACS, Embase, CINAHL, Scopus, WoS, CARPHA; Mendeley reference manager. A random-effects meta-analysis model was applied, and heterogeneity was assessed using the Higgins test. **Results**: Out of 41 studies included in the review, involving a total of 93,461 patients with leprosy, 4.09% (n=3,830) were eligible for relapse after polychemotherapy. Of them, 69.71% (n=2,670) were treated both with multibacillary and with paucibacillary regimes (72.36%, n=1,932; and 27.64%, n=738, respectively), and with a bacilloscopy index ≥4. Relapse prevalence was observed in males and in people aged over 30 years old. The meta-analysis estimated the global prevalence of leprosy relapse at 11% (95%CI: 0.09-0.12), with higher prevalence rates in Brazil (31%) and India (13%). **Conclusion**: There is evidence of high global prevalence of leprosy relapse after PCT, with higher estimates in India and Brazil, countries burdened with higher prevalence of the disease.

**SYNTHESIS:** Although leprosy us an ancient disease with a scientifically proven effective treatment, it remains a Public Health problem. This is not only due to the disease high prevalence but also to its potential to cause physical disabilities, leading to emotional and social impacts and, consequently, compromising quality of life. In addition to the new cases of the disease, another concern commonly reported in the literature is leprosy relapse after polychemotherapy, as it has repercussions on therapeutic effectiveness. The relapse causes are usually associated with therapeutic failure due to incomplete treatment, misclassification in the initial treatment, and multidrug resistance. This study provides insights to verify the disease current prevalence based on scientific evidence, which can contribute to expanding the prevention strategies.

## INTRODUCTION

Leprosy is a chronic infectious disease of a dermatoneurological nature, caused by *Mycobacterium leprae* or by *M. lepromatosis. M. leprae* presents tropism for skin cells and peripheral nerves, potentially causing deformities and physical disabilities that exert impacts on the social, emotional and psychological aspects of the patients’ lives, adversely affecting their quality of life^1, 2^.

Since 1981, leprosy has been treated with the Polychemotherapy (PCT) regime recommended by the World Health Organization (WHO), aimed at eliminating active bacillus transmission within communities^3,4^. Currently, leprosy patients receive a combination of three drugs during treatment, regardless of the clinical form, with only the number of doses differing. Paucibacillary (PB) patients are administered clofazimine, dapsone and rifampicin in a 6-dose regime, whereas Multibacillary (MB) patients undergo a 12-dose regime. Although PCT has been an important tool in leprosy control, therapeutic regime failures, usually caused by diagnostic errors, inadequate treatment periods and irregular medication intake, can compromise leprosy surveillance effectiveness and trigger disease relapse^5–6^. Despite being uncommon when compared to other pathogens, we must not forget the existence of resistant strains.

Leprosy relapse is characterized by reoccurrence of the disease clinical activity (cutaneous and/or neurological) after regular treatment with standardized regimes, that is, when the patient is discharged as cured^3,7,8,9,10^. Leprosy relapse cases typically occur between 2 and 15 years after having concluded the PCT treatment^11^. According to the operational classification, the time for relapse occurrence is commonly seen in PB patients within five years after PCT and in MB patients in an equal or longer period of time^12–17^.

In 2021, it was estimated that 106 countries reported 140,594 new cases of the disease, indicating a 10.2% increase in the detection rate of new cases when compared to 2020. Also in 2021 there were 15,516 retreatment cases due to leprosy relapse, with 20% (3,201 cases) in 51 countries reporting relapse in patients after having concluded PCT, with the highest numbers in Brazil (1,212; 37%) and India (510; 16%)^18–22^.

With the advent of the COVID-19 pandemic, a global 37.1% reduction in newly recorded cases was observed when comparing 2020 (n=127,396) to 2019 (n=202,488). This suggests underreporting of cases in these years, which may also impact relapse cases ^23^.

Another issue that may be linked to leprosy relapse is antimicrobial resistance. WHO data on MDT resistance indicate that, out of 3,452 tested patients, 51 (1.5%) were diagnosed with *M. leprae* resistant to rifampicin, 49 (0.23%) to dapsone, three (0.1%) to ofloxacin, and four (0.11%) presented strains resistant to more than one antimicrobial^18–22^.

Considering that, although relapse cases are not included in the disease incidence indicator, they influence prevalence of the disease. In addition, the severity of leprosy relapse is not only related to bacillus infection but also to the social, emotional and psychological impacts involved in the illness context and people’s experiences with the disease (such as long treatment periods, leprosy reactions, pain, stigma and disability).

It is in this context that the current systematic review becomes relevant. A preliminary search conducted on PROSPERO, MEDLINE, the Cochrane Database of Systematic Reviews and JBI Evidence Synthesis did not identify any current or ongoing systematic reviews on the theme.

This study aimed at synthesizing the best scientific evidence related to estimating the prevalence of leprosy relapse cases after polychemotherapy. Such knowledge might contribute more robust information on the topic, aiding in the planning of strategies for clinical management and surveillance in the care practice focused on the disease.

### Review questions

Which is the estimated prevalence of leprosy relapse after PCT? Which are the characteristics of the studies related to clinical specificity (clinical form, relapse occurrence timing, bacilloscopy, histopathology, physical disability degree and multidrug resistance to the PCT components)? Which is the global estimate, by geographical region, of the relapse prevalence in the cases studied?

### Inclusion criteria

The inclusion criteria were defined based on the PopCoCo mnemonic (Population, Condition, and Context). **Pop**: patients of any age and gender diagnosed with leprosy relapse after regular PCT treatment. **Co**: leprosy relapse was defined as patients who, within a specific period, were diagnosed with retreatment due to relapse, meaning that they presented clinical activity of the disease (cutaneous and/or neurological) after regular treatment with a standardized regime (PB and MB), that is, they were discharged as cured. Relapse prevalence (%) was estimated (combined global prevalence and its respective confidence interval [CI]) by the countries where the primary studies were conducted. Relapse (number and %) concerning clinical specificity (PB/MB, relapse occurrence timing, bacilloscopy, physical disability degree [PDD], histopathology and multidrug resistance to PCT). **Co**: considered for any geographical region and health care level where the patient was diagnosed or underwent treatment.

### Types of study

This review considered observational studies, such as analytical cross-sectional, retrospective and prospective cohort, case-control and experimental studies.

Studies lacking the frequency of relapse cases in the study population were excluded, as well as those where relapse was due to abandonment, irregular treatment, monotherapy or alternative treatment.

### Method

This is a systematic review of prevalence studies following the JBI methodology^24,25^. The systematic review protocol was registered in the International Prospective Register of Systematic Reviews (PROSPERO: CRD42020177141). The reporting stage adhered to the PRISMA recommendations.

### Search strategy

An initial search limited to MEDLINE was conducted using Medical Subject Headings (MeSH) and related keywords (leprosy OR “Mycobacterium leprae” OR “relapse” OR “risk factors” OR prevalence).. This search was followed by a word analysis in the titles, abstracts and indexing terms used to describe the studies.

A second search, using all the keywords and indexing terms identified, was performed in the following databases: National Center for Biotechnology Information (NCBI) from the National Library of Medicine (NLM)/ Medical Literature Analysis and Retrieval System Online (PubMed/MEDLINE); *Literatura Latino-Americana e do Caribe em Ciências da Saúde* (LILACS); Embase; Cumulative Index to Nursing and Allied Health Literature (CINAHL); Scopus; Web of Science (WoS); and, finally, the Caribbean Public Health Agency (CARPHA) for unpublished studies (Gray Literature). In the third stage, the reference lists of the studies selected in the full-reading phase were accessed.

The MeSH index term search included relapse, leprosy, risk factors and prevalence. The studies were identified based on a time clipping at 1981 due to the implementation of the treatment recommended by the WHO.

A preliminary search in the PubMed/Medline database was conducted ((((leprosy[MeSH Terms]) OR (mycobacterium leprae[MeSH Terms])) OR (multibacillary leprosy[MeSH Terms])) OR (paucibacillary leprosy[MeSH Terms])) AND (((((relapse[MeSH Terms]) OR (recurrence[MeSH Terms])) OR (risk factors[MeSH Terms])) OR (determinants health[MeSH Terms])) OR (prevalence[MeSH Terms])). From this strategy, other search strategies were developed according to each database.

### Selection of the studies

All the studies identified were selected by two reviewers and exported in RIS (Research Information Systems) format to the Mendeley Desktop reference manager, version 1803, followed by SUMARI JBI24^24^.

### Methodological quality assessment

The critical evaluation of the studies’ methodological quality was conducted by two independent reviewers. In cases of assessments that resulted in disagreement, a third reviewer was consulted for evaluation. The quality of the publications included was assessed based on criteria derived from the JBI critical appraisal checklist for observational and experimental studies^24^.

The cutoff point to include a study in the review was having 50% or more “Yes” answers to the standardized questions from the JBI critical appraisal tool for studies containing prevalence data, ensuring greater confidence in the methodological rigor of the method used.

### Data extraction

Data extraction was performed by independent reviewers. A data extraction table was used, created to assess the quality of demographic data, study locus, sample size, number of leprosy and relapse cases, number of PB and MB cases in the initial treatment and relapse, following the essential information from the JBI SUMARI data extraction instrument for systematic reviews of prevalence studies^24^.

### Data synthesis

The analyses were conducted in the Stata software, version 13.1.

Observational cohort studies were included for the meta-analysis. The inclusion of homogeneous studies enabled a meta-analysis of the relapse prevalence (%), estimating the combined global relapse prevalence and by country where each primary study was conducted. The random-effects meta-analysis model was applied to detail the overall combined relapse prevalence (global estimate). Heterogeneity across the studies was analyzed using the Higgins test (I²), which indicates the variation percentage between studies through Confidence Intervals (95%CIs).

## RESULTS

### Study selection process

The search resulted in a total of 7,814 studies identified in the databases and five records from other reference sources. Of these, 5,216 studies were excluded for being duplicates, and 2,603 records were selected to read their titles and abstracts. Next, 107 studies were chosen for full-text reading and subsequent methodological assessment. A total of 41 studies were included in the review, and 18 in the meta-analysis, as presented in the Preferred Reporting Items for Systematic Reviews and Meta-Analyses, JBI (Figure 1).

**Figure 1.**
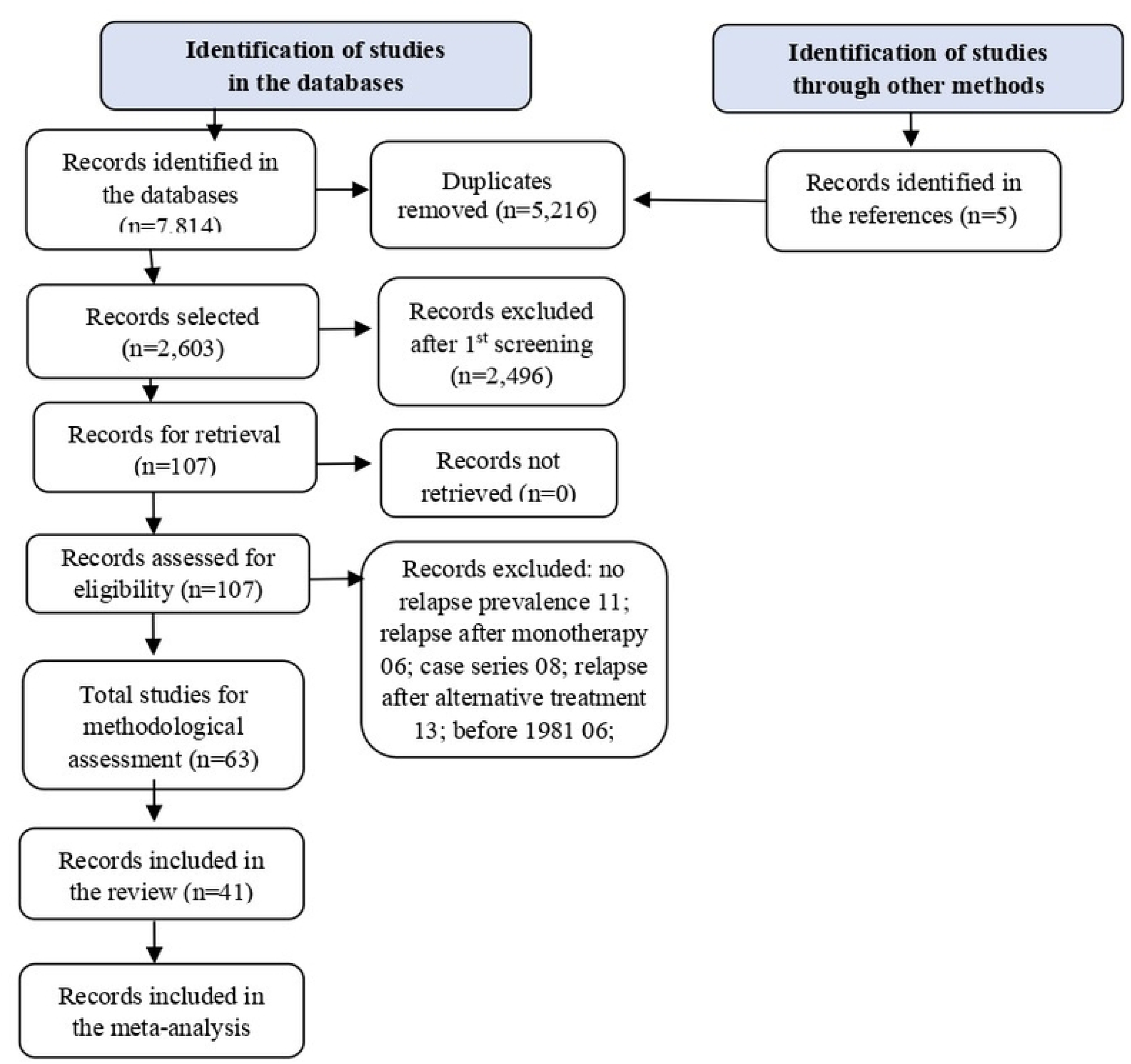
Synthesis of the systematic review stages according to the Preferred Reporting Items for Systematic Reviews and Meta-Analyses^(25)(24)^.

### Characteristics of the population and clinical aspects according to the study designs

Of the 41 studies included in the review, totaling 93,461 leprosy patients, 3,830 (4.09%) were eligible for relapse after PCT. Within them, 2,670 (69.71%) underwent MB regimes (1,932; 72.36%) and PB regimes (738; 27.64%).

The main findings from the studies indicated that the relapse occurrence time after the initial PCT treatment is equal to or greater than 5 years after clinical cure, with higher prevalence of patients treated for MB^26,27,28,29^. Complementary findings from the cohort studies revealed that relapse cases were more prevalent among cases with positive bacilloscopy when compared to those with negative bacilloscopy in the initial treatment^13,14,15,30,31,32,33,34,35,36–50^.

The characteristics of these 18 studies included in the meta-analysis are presented in Chart 1^14,15,31,32,33,34,36,37,39,40,42,45,46,47,48,49,50,51^. Of the total population suffering from leprosy analyzed in the meta-analysis (n=74,742), 867 (1.15%) were treated as relapse. Eight (0.93%) of these studies were classified as MB relapse^14,15,32,34,42,46,47,51^ and four (0.58%) by the PB classification^15,32,46,50,51^. In the 12 studies, the male gender was more prevalent^14,31,32,34,36,37,39,40,42,45,46,48^ and the mean age was over 30 years old, with one study having a mean age below 15 years old^14,15,31,32,34,36,37,42,45,46^. A bacilloscopy index ≥4 BI was observed in the studies evaluated^14,31,32,33,34,36,37,40,42,45,46^. Histopathology was assessed in twelve studies, with prevalence of the borderline-lepromatous and lepromatous clinical forms^14,31,32,34,36,37,40,42,45,49,48,49^. PDD was analyzed in seven studies, with prevalence of grade 1^14,15,31,32,37,40,51^. Among the studies evaluated for information on resistance to PCT, one study reported single resistance to rifampicin, dapsone and resistance to rifampicin + dapsone^31^.

**Chart 01-.**
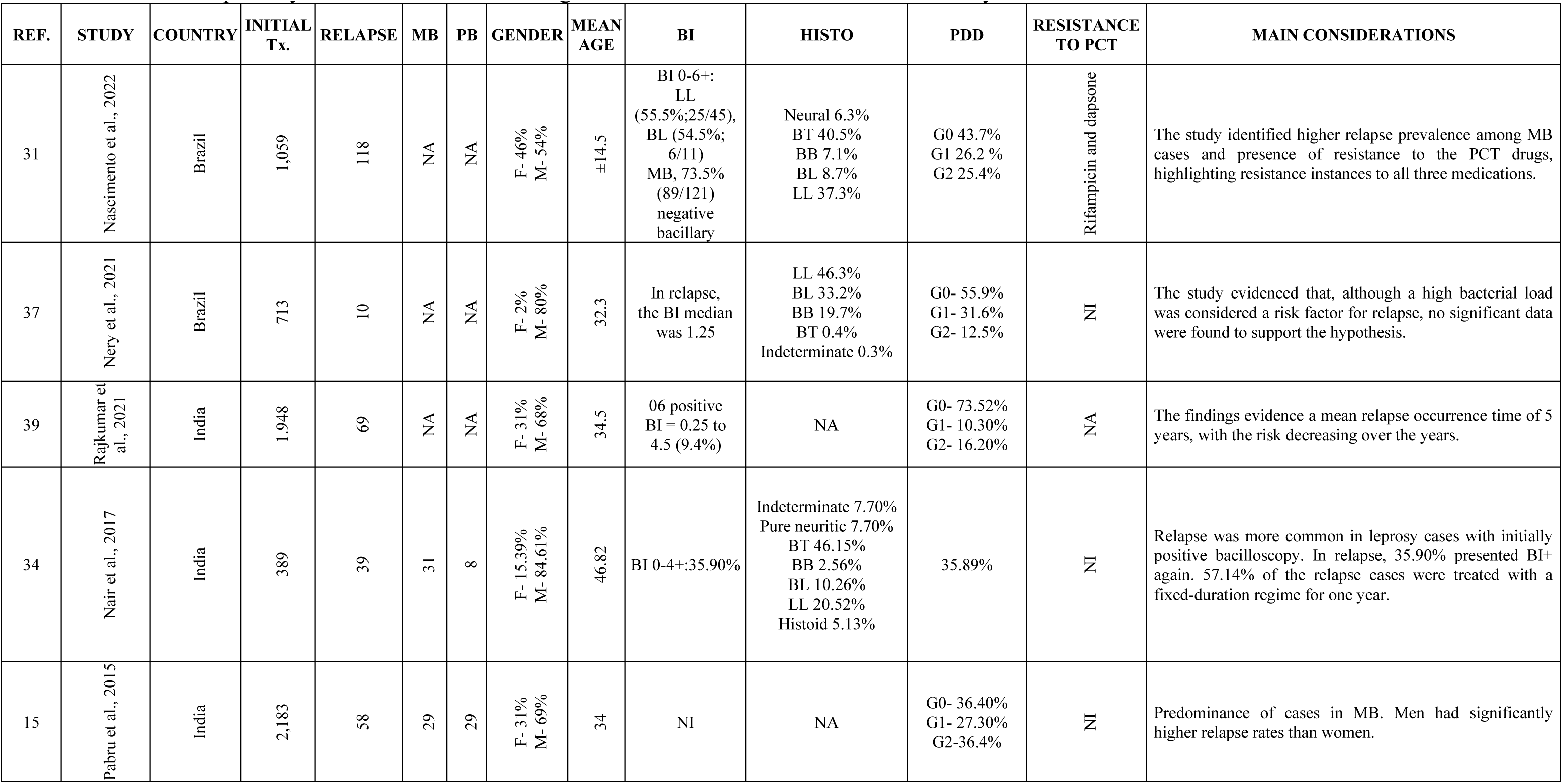
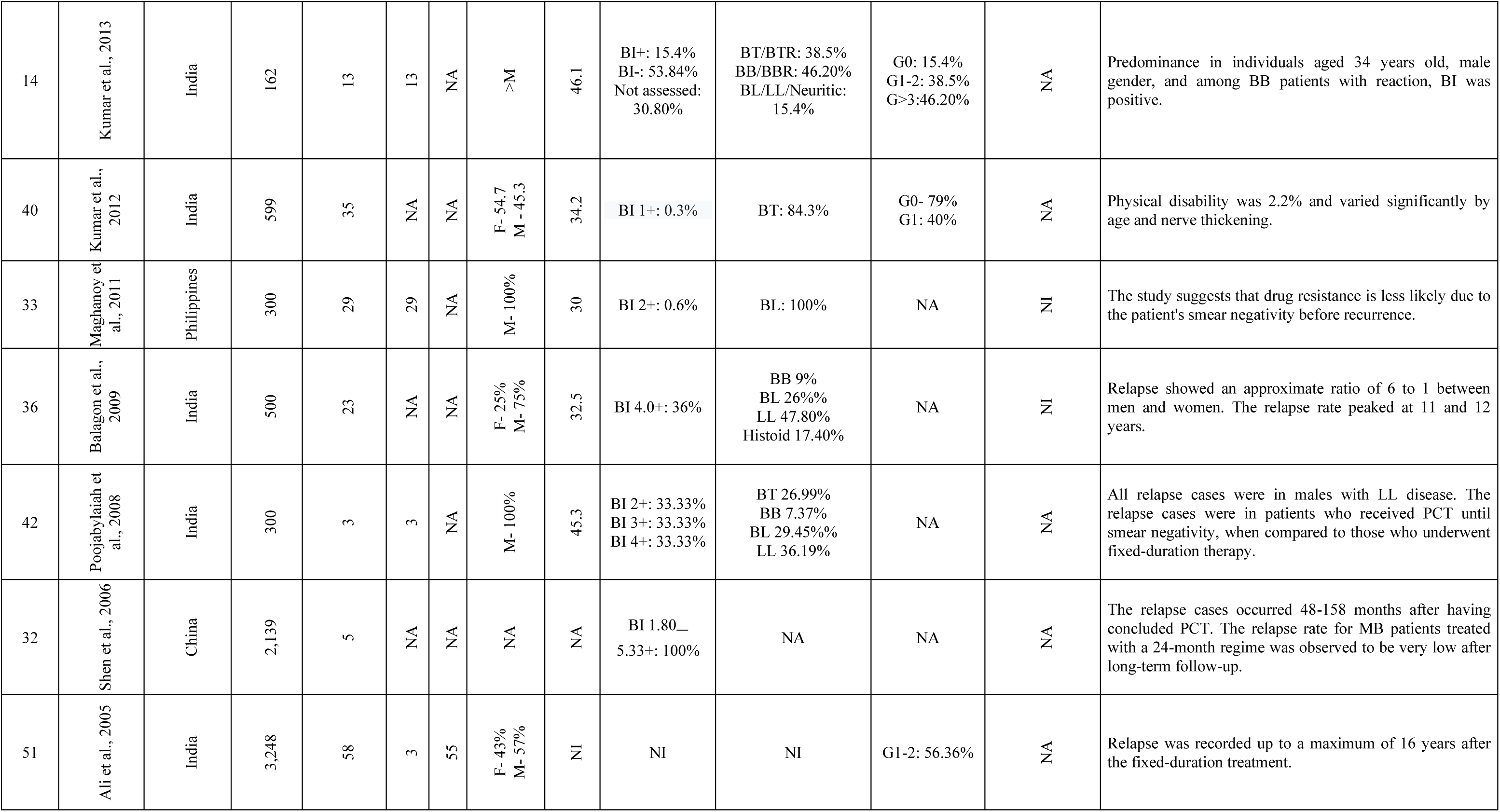
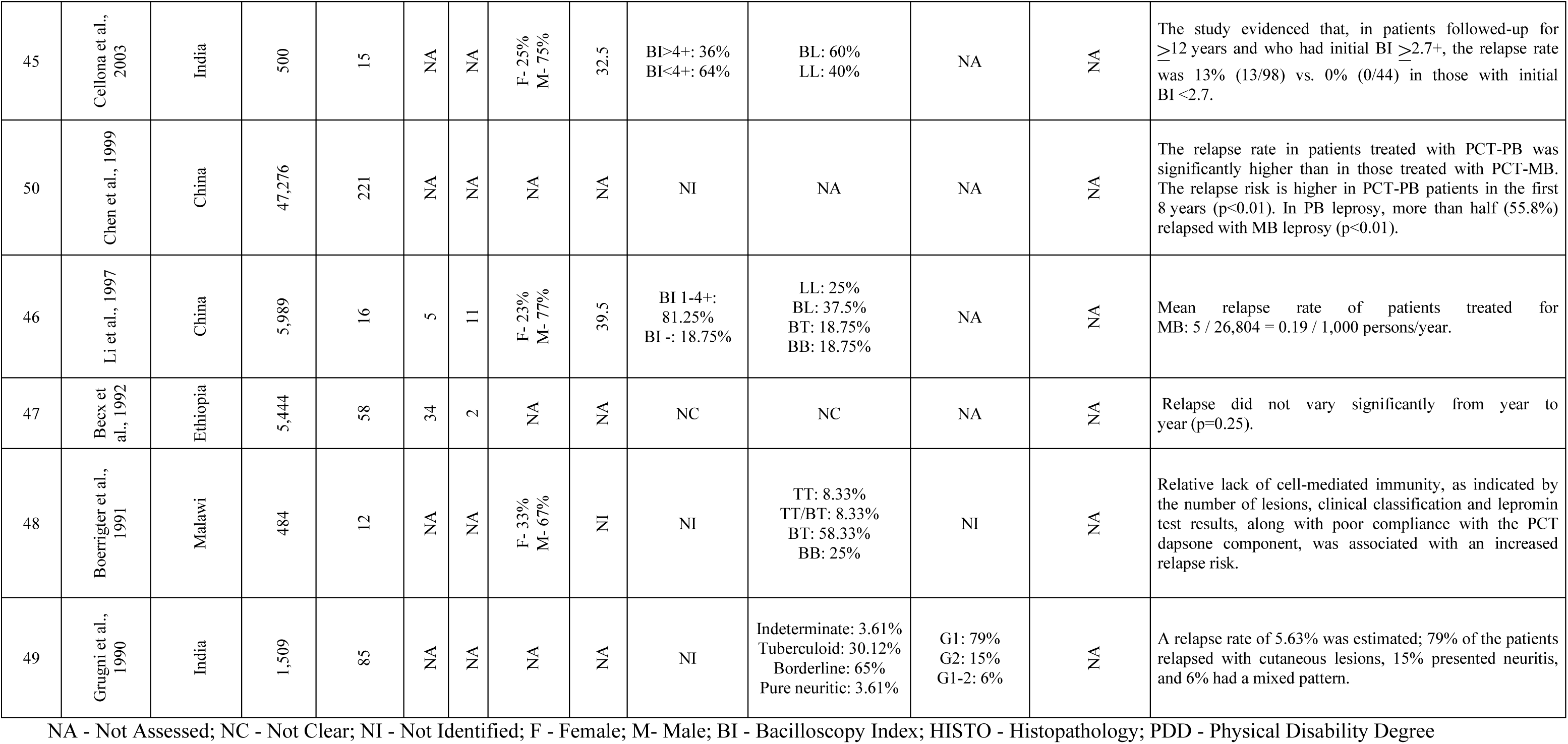
Descriptive synthesis of the main findings of the studies included in the meta-analysis.

### Meta-analysis: Synthesis of the prevalence review findings

The overall prevalence of leprosy relapse was 11% (95%CI: 0.090-0.12) with a null effect. Heterogeneity was assessed using the I^2^ statistic and was very high (>90%). Among the eight studies with the largest populations, individually assessed, the prevalence of relapse varied from 0% to 0.06% (weight: 5.27-5.49). The subgroup of studies conducted in China indicated 0% prevalence (95%CI: 0.00, 0.01). Two countries presented higher relapse prevalence percentages, with Brazil at 31% (95%CI: 0.07, 0.56), followed by India at 13% (95%CI: 0.05, 0.20), as shown in Table 1 and Figure 02.

**Table 1-.** Synthesis of the data from the meta-analysis studies.

| REF. | STUDY | COUNTRY | TOTAL POP. | RELAPSE POP. | WEIGHT | 95%CI |
| --- | --- | --- | --- | --- | --- | --- |
| 31 | Nascimento et al., 2022 | Brazil | 1,059 | 118 | 4.94 | 0.09, 0.13 |
| 37 | Nery et al., 2021 | Brazil | 713 | 10 | 5.37 | 0.01, 0.03 |
| 39 | Rajkumar et al., 2021 | India | 1,948 | 69 | 5.38 | 0.03, 0.04 |
| 34 | Nair et al., 2017 | India | 389 | 39 | 4.30 | 0.07, 0.13 |
| 15 | Pabru et al., 2015 | India | 2,183 | 58 | 5.42 | 0.02, 0.03 |
| 14 | Kumar et al., 2013 | India | 162 | 13 | 3.55 | 0.05, 0.13 |
| 40 | Kumar et al., 2012 | India | 599 | 35 | 4.95 | 0.04, 0.08 |
| 33 | Maghanoy et al., 2011 | Philippines | 300 | 29 | 4.07 | 0.07, 0.14 |
| 36 | Balagon et al., 2009 | India | 500 | 23 | 4.97 | 0.03, 0.07 |
| 42 | Poojabylaiah et al., 2008 | India | 300 | 3 | 5.28 | 0.00, 0.03 |
| 32 | Shen et al., 2006 | China | 2,139 | 5 | 5.49 | 0.00, 0.01 |
| 51 | Ali et al., 2005 | India | 3,248 | 58 | 5.46 | 0.01, 0.02 |
| 45 | Cellona et al., 2003 | India | 500 | 15 | 5.13 | 0.02, 0.05 |
| 50 | Chen et al., 1999 | China | 47,276 | 221 | 5.49 | 0.00, 0.01 |
| 46 | Li et al., 1997 | China | 5,989 | 16 | 5.49 | 0.00, 0.00 |
| 47 | Becx et al., 1992 | Ethiopia | 5,444 | 58 | 5.48 | 0.01, 0.01 |
| 48 | Boerrigter et al., 1991 | Malawi | 484 | 12 | 5.18 | 0.01, 0.04 |
| 49 | Grugni et al., 1990 | India | 1,509 | 85 | 5.27 | 0.05, 0.07 |
| **Total** |  |  | **74,742** | **867** | **100.00** | **0.09, 0.12** |

**Figure 02.**
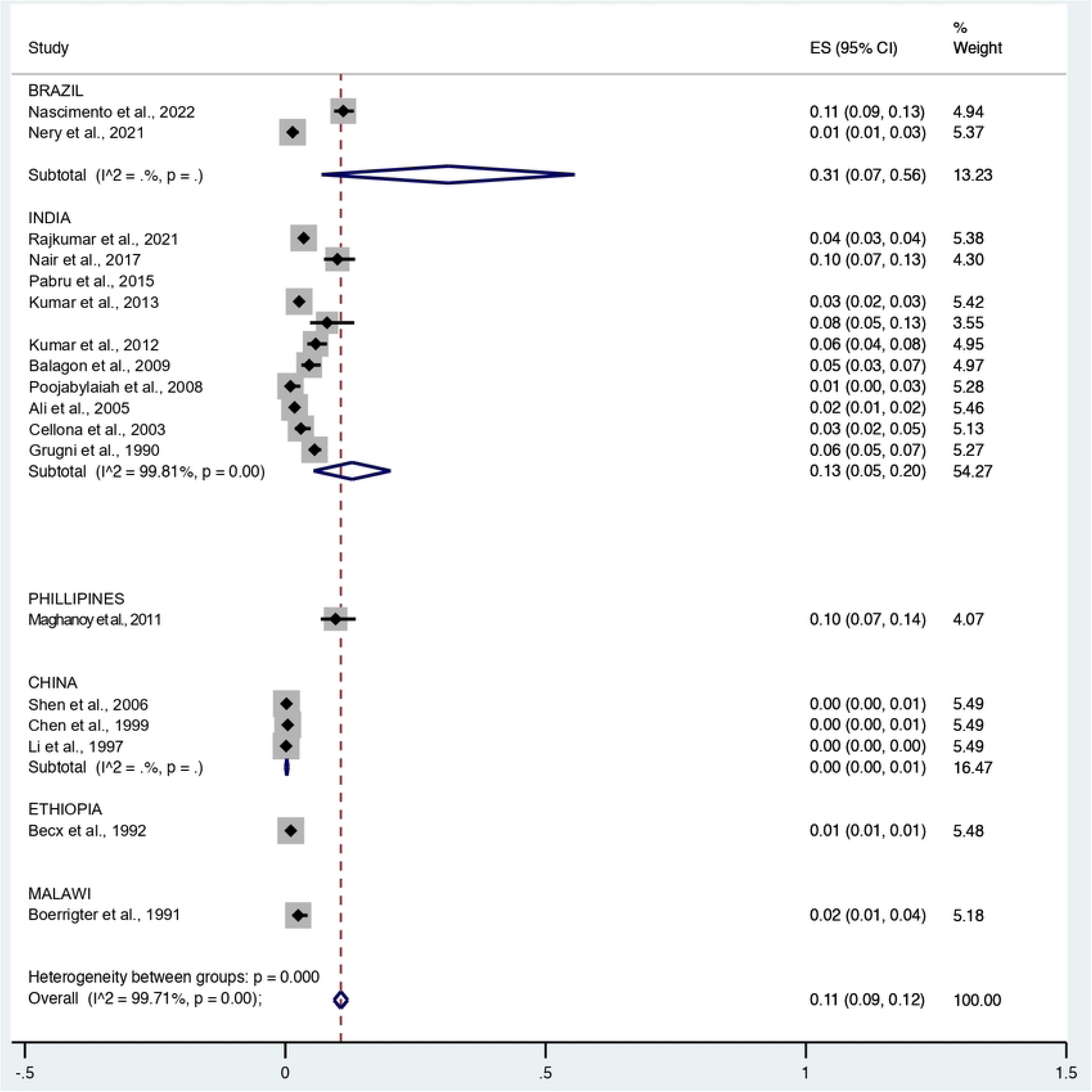
Forest plot for a random-effects meta-analysis of the overall leprosy relapse prevalence by country where each study was conducted.

## DISCUSSION

The clinical characteristics of the studies included showed that the relapse occurrence time was consistently equal to or greater than 5 years after clinical cure. In diagnosing relapse, higher prevalence was observed among male patients aged over 30 years old. There was relapse predominance in multibacillary patients with positive bacilloscopy and the borderline tuberculoid and lepromatous relapse clinical forms in patients with grade 1 physical disabilities. The systematic review also indicated multidrug resistance to PCT, with drug-resistant variants related to single resistance to rifampicin and dapsone, as well as resistance to rifampicin + dapsone.

The regions analyzed in this review included India, Brazil, Indonesia, China, Malawi, Ethiopia, Colombia and the Philippines. The global prevalence estimate in the studies analyzed in the meta-analysis was 11%, although it presented a null effect. When evaluating the subgroups based on the country where each study was conducted, Brazil had the highest prevalence of cases (31%), followed by India (13%).

In our study, the systematic review of prevalence data is important for describing the geographical distribution among subgroups, providing information for health care planning and resource allocation. Therefore, it is necessary to monitor the trends in burden and emergence of the disease, in this case, the relapse prevalence. The countries historically reporting the highest numbers of newly detected cases worldwide are India and Brazil^20,23^. Therefore, planning guidelines should prioritize to minimize the disease burden in these regions, focusing on systematic care for early and timely treatment of all cases and systematic surveillance of the contacts^53,54^.

The primary indicator of treatment efficacy in leprosy cases is the low relapse percentage after its conclusion. However, relapse cases may be underestimated. Therefore, some studies suggest that occurrences of acute inflammatory episodes corresponding to leprosy reactions after therapeutic treatment conclusion can be frequent in the early years after treatment. However, they need to be differentiated from relapses, requiring clinical and laboratory resources^55,56^. The findings also suggest that countless aspects related to the disease lack more robust primary studies, mainly regarding factors associated with relapse occurrence in the population, particularly in countries with higher burden of the disease.

The meta-analysis also evidenced that, among the eight studies with a larger population evaluated individually, the prevalence of relapse varied from 0.00% to 0.06%. Considering that the studies conducted in China showed 0% prevalence, it also suggests more precise and methodologically robust studies. Despite a reduction in the relapse prevalence is noticed in these groups, the magnitude and high disabling power of the disease continue to make it a Public Health problem^23,57^.

According to WHO data in 2021, 20% of 51 countries reported retreatment due to relapse after MDT, with a higher proportion in Brazil (37%) and India (16%). This is in line with the higher prevalence findings obtained in this review for these countries. Although the meta-analysis indicates a null effect of overall relapse, it is crucial to prioritize leprosy control strategies mainly in these countries. The relapse incidence has a multifactorial context but. in some cases, it may indicate therapeutic failure. In addition, the prevalence of relapse cases with positive bacilloscopy poses a significant obstacle, as they are considered “active” infectious cases capable of transmitting leprosy^35,38,58,59^.

Therefore, implementing early and accurate diagnosis and treatment, as well as eliminating active sources of infection, is one of the main challenges in reducing the burden of this disease. In this context, the WHO operational classification system based on the number of skin lesions can mainly lead to misclassification of MB as PB cases, consequently increasing the relapse chances in the more severe forms of the disease^50,61^.

In this review, the studies evidence higher occurrence of leprosy relapse in a period equal to or more than 5 years after PCT treatment. This suggests the need for improvements in clinical management strategies and to expand the systematic surveillance period for cases that have been discharged as cured in health services^12,27,33,62^.

The higher occurrence of relapse in men aged at least 30 years old can be associated with the fact that they are more affected as new cases, low health service utilization, environmental factors in the person’s occupation, or men’s hormonal predisposition to the bacillus, as highlighted in some studies^12,29,63,64,65^.

A higher relapse prevalence was observed among the MB leprosy cases. Patients with MB leprosy are at a higher risk of developing relapse when compared to those with PB leprosy, which can be related to the fact that MB patients have high bacillary loads. The importance of each person’s genetics cannot be overlooked^31,52^. However, it is worth noting that, among the primary studies classified as PB in the first treatment, there was higher relapse prevalence as MB, suggesting potential errors in the disease operational classification in the initial treatment ^32.66,^

Some studies estimate that the chance of correctly categorizing an MB patient is 71.3%, turning diagnostic errors into one of the main factors associated with disease relapse^62,67^. In addition to increasing the relapse chances, this contributes to persistent presence of bacilli in the communities, as indicated by results from genetic sequencing studies that identified presence of MDT-resistant bacilli in the initial treatment, and more frequently among the relapse cases^34,35, 67,68,69^. The findings also emphasize that people living in endemic and hyperendemic areas have natural susceptibility to infection and relapse due to endogenous or exogenous reinfection by the *M. leprae*^53,54,55,62,65^ bacillus.

The reinfection issue in these cases can be related not only to individual factors such as immunological deficiency to the bacillus but mainly to the deficit in strategies for the epidemiological surveillance of cases and contacts. There is a need to invest in *M. leprae* detection through molecular biology techniques, such as laboratory support tools^34,61,66,67^.

Currently, the diagnostic methods available in health services are mostly bacilloscopy, histopathology and the Mitsuda test, which alone are not sensitive or specific for the correct clinical and operational classification of leprosy, with the possibility of leading to misclassification of MB as PB cases. The results of these tests can increase the relapse chances in more severe forms of the disease^55,60^.

Expanding access to diagnostic tests that assist in the correct operational classification in the initial treatment is essential for controlling spread of the bacilli, especially the resistant ones, in regions considered hyperendemic for the disease, such as diagnostic tests based on molecular techniques using biomarkers like PCR and NDO-LID^55,60,62,68,69,70^.

Regarding resistance to PCT, although only one study^33^ from those included in the meta-analysis analyzed this factor associated with relapse, other studies in the review, not included in the meta-analysis, provided data on mutations in the *M. leprae* genes, presenting resistance to dapsone and rifampin. In addition, patients with mutations for resistance to dapsone and rifampin and genes with mutations resistant to all three PCT drugs were reported in several studies^35,53,54,70, 71,72,73,74,75^.

Some studies reinforce that global efforts to control leprosy through intensive chemotherapy have led to a significant decrease in the number of patients recorded; however, it is indispensable to monitor for multidrug resistance for maintaining treatment effectiveness. Studies conducted in hyperendemic countries have shown multidrug resistance to rifampin and dapsone in new cases and relapses. When combined with molecular resistance and VNTR, some data revealed diverse evidence of intrafamilial primary transmission of resistant *M. leprae*^35,54^.

Thus, in a context of controlling spread of the bacillus, correct and early diagnosis and extended surveillance of cured cases and contacts are crucial for reducing the number of new cases and relapse of the disease^3,6,17,22,37,46,76^. The unified treatment (PCT-U) that would waive the need for operational classification, recommended in 2020 by the Ministry of Health in Brazil, is not yet a national or global reality and was not analyzed in this review^62, 63,64,72,78,79^.

Therefore, the results of this review reinforce the need for systematic follow-up of leprosy-treated patients, aimed at screening reactivation cases due to relapse of the disease, which can cause reinfections resistant to the standardized regimes.

### Limitations

This review presented some limitations, as not all the studies included provided information regarding the operational classification at the initial diagnosis and at the relapse time, making it impossible to perform a meta-analysis regarding the disease relapse operational classification. The limited information on age and gender in the primary studies also posed hindered the analyses. Nevertheless, the conclusions drawn from the analysis of the studies included can contribute to expanding the available knowledge and serve as evidence to aid decision-making regarding control of the disease, especially in the current context of introducing PCT-U.

## CONCLUSION

The systematic review with meta-analysis indicated high global prevalence of leprosy relapse after PCT, with higher estimates in India and Brazil, countries with greater burdens of the disease.

### Recommendations for the clinical practice

Based on the findings of this review, it is recommended to formulate Public Health policies to: ground and guide care for people with leprosy and their contacts; expand access to diagnostic tests both for initial diagnosis and for early relapse: invest in health professionals’ training to improve systematic monitoring of cases and contacts; as well as develop more universally effective strategies, mainly in countries such as India and Brazil, which presented a higher global estimated prevalence of leprosy relapse.

In addition, the findings suggest the need for robust studies on the leprosy bacillus resistance to the PCT drugs among new leprosy cases and to develop diagnostic methods to detect this resistance and reinfection cases.

### Recommendation for research

Based on this review, there is a notable need for expanding molecular studies aimed at accurately detecting resistant strains and reinfection cases. These studies might provide crucial information for decision-making regarding future chemotherapeutic approaches in leprosy control, particularly given the current scenario and the need to address problems related to diagnostic errors in the clinical condition, mainly in the current model of the unified treatment (PCT-U) for leprosy. Therefore, they might contribute to the implementation of more assertive Public Health policies and surveillance strategies, ultimately reducing the socioeconomic impact caused by the disease.

Some factors require more robust studies, especially those associated with leprosy relapse occurrence since, despite the reduction in new cases, there is still significant prevalence of leprosy relapse cases in the population, especially in countries with higher burdens of the disease.

## Data Availability

All relevant data are in the manuscript and its supporting information files.

